# Witnessing Trauma in the Modern Era: The Role of Uncensored Media in Mental Health

**DOI:** 10.64898/2026.03.16.26348519

**Authors:** Hadas Allouche-Kam, Tal Elhasid Felsenstein, Isha H Arora, Alon Bartal, Christina T Pham, Sabrina J Chan, Sharon Dekel

**Affiliations:** Postpartum Traumatic Stress Disorders Research Program, Department of Psychiatry, Massachusetts General Hospital, Department of Psychiatry, Mass General Brigham, Boston, MA; Department of Psychiatry, Harvard Medical School, Department of Psychiatry, Boston, MA; School of Business Administration, Bar-Ilan University, Ramat Gan, Israel

**Keywords:** Digital media exposure, Uncensored traumatic content, Peripartum mental health, Post-traumatic stress symptoms, PTSD, Collective trauma, Social media

## Abstract

**Background:** Digital media increasingly shape how populations encounter large-scale traumatic events, enabling real-time exposure to uncensored graphic content among individuals who are not directly exposed. However, whether this form of indirect exposure to the trauma relates to posttraumatic stress responses, particularly in the wake of collective, large-scale trauma, remains poorly understood.

**Methods:** We studied a large cohort of individuals in the first months following a collective trauma, in which a significant portion reported symptoms of post-traumatic stress disorder (PTSD) related to the October 7th events in 2023 although none were directly exposed. Participants were assessed for mental health symptoms, demographic background, social and psychological factors, and degree of trauma exposure concerning geographic, i.e., physical proximity from threat, interpersonal, e, g., death of close family/friend, and media, i.e., censored and uncensored watching and reading trauma content.

**Results:** Around 24% of the sample met clinical threshold for PTSD. Intrusive and hyperarousal symptom clusters were commonly endorsed. Hierarchical regression analysis revealed that greater exposure to uncensored traumatic video content through affected social networks was associated with higher PTSD symptom severity, above and beyond other important risk factors including mental health history, reduced perceived resilience and social support, and degree of religiosity, and other forms of trauma exposure.

**Conclusions:** The findings identify exposure to uncensored traumatic digital content as a distinct dimension of indirect trauma exposure and suggest that features of contemporary media environments may shape early post-traumatic responses during collective crises.

## 1. Introduction

Collective trauma refers to the shared psychological impact of large-scale events involving mass violence or threat that extends beyond individuals directly exposed to the events (Hirschberger, 2018). In recent years, collective trauma has increasingly been circulated through social media, embedding graphic depictions of distant violence into everyday perception and creating psychological proximity to unfolding events for non-firsthand exposed individuals.

This evolving form of experiencing and witnessing traumatic events warrants closer examination of exposure definitions within diagnostic frameworks. According to the *Diagnostic and Statistical Manual of Mental Disorders (*5th ed*)(American Psychiatric Association, 2013)*, Qualifying exposure criteria for a traumatic event that may result in post-traumatic stress disorder (PTSD) include direct personal experience of the event, firsthand witnessing of the trauma occurring to others, indirect exposure through learning that the event occurred to close others, or repeated occupational exposure to traumatic material. While media exposure can function as a form of vicarious trauma, particularly when content is graphic, personalized, or identity-relevant, DSM-5 explicitly excludes non-occupational media exposure. It warrants investigation to better understand whether media exposure to large-scale violent events such as collective trauma can fuel clinically significant psychological distress in individuals outside those directly affected.

Experimental work reveals short-term stress responses to media exposure (James et al., 2016). More importantly, observational studies in naturalistic settings, such as in the aftermath of the September 11 attacks and Boston Marathon bombings, suggest that repeated exposure to widely shared images of violence can result in acute stress and arousal among individuals geographically distant from the event, with evidence, although mixed, in support of persistent symptoms (Ahern et al., 2002; Holman et al., 2014; Silver et al., 2002). While media exposure could be regarded as a causal driver, it may merely signify a marker of distress among individuals exposed to the trauma through other forms of indirect exposure or those with pre-existing mental health problems.

The impact of media on traumatic stress responses to collective trauma has gained renewed relevance in the digital era, as social-media platforms enable widespread exposure to uncensored traumatic content. Unlike traditional broadcast media, in which violent content is edited and time-limited, and embedded within structured journalistic narratives, digital environments allow real-time, visually intense, and repeated exposure to violent material (Lamba et al., 2023; Silver et al., 2013). This form of “digital witnessing” allows individuals not directly exposed to vividly observe a traumatic event as it unfolds (Holman et al., 2020). Although social media exposure may heighten stress responses and vicarious trauma, research linking such exposure to clinically significant PTSD is limited, primarily in the context of collective traumatic events, with little evidence from naturalistic settings (Abdalla et al., 2021; Hall et al., 2019; Neria and Sullivan, 2011). To the best of our knowledge, whether violent exposure via media independently contributes to the endorsement of PTSD symptoms in the wake of collective trauma is unknown.

To this end, we studied a large cohort of individuals in the aftermath of a collective traumatic event. None had direct exposure, but some developed symptoms of PTSD. We examined the unique contribution of media exposure to endorsement of PTSD symptoms, with a focus on censored and uncensored violent media content. Specifically, we tested the independent role of media exposure above and beyond known PTSD predictors and other forms of indirect exposure.

## 2. Methods

### 2.1 Participants

The present study is part of a research investigation concerning the psychological sequelae of October 7, 2023 terrorist attacks and the Israel–Hamas War on peripartum women residing in Israel (Allouche-Kam et al., 2025). This study included two waves of data collection conducted several months apart. The first wave was collected between January and March 2024, during the war. Participants aged 18 years or older who were pregnant or within six months postpartum and resided in Israel during the war completed questionnaires assessing sociodemographic background, mental health and related factors, psychosocial resources, and trauma-exposure. The study was approved by the Mass General Brigham Human Research Committee. Participants provided implied consent by responding to study questionnaires.

The sample in this study consisted of 630 women who provided information regarding media exposure, excluding participants (n=17) with direct exposure to the attacks (i.e., endorsed “I survived the massacre”). Mean age of participants was 32 (Mean= 32.7, SD = 4.4). The majority were married (98.6%), had attained at least a bachelor’s degree (90.5%), and were Jewish (98.9%). Around half (57%) of participants were postpartum. Detailed demographic and clinical characteristics are presented in Table 1.

**Table 1.** Sociodemographic, Reproductive, and Trauma-Related Characteristics of the Study.

| *Variable* | *n (%) / M (SD)* |
| --- | --- |
| **Demographic Factors** |  |
| Maternal Age | 32.73 ± 4.42 |
| History of mental illness | 71 (11.27%) |
| Marital Status |  |
| Married or domestic partnership | 621 (98.57%) |
| Single, not in a relationship | 9 (1.43%) |
| Education |  |
| Less than a bachelor's degree | 60 (9.52%) |
| Bachelor's Degree | 306 (48.57%) |
| Graduate degree (Master's/Doctoral/Medical) | 264 (41.90%) |
| Religiosity |  |
| Very to slightly religious | 289 (45.87%) |
| Not religious at all | 341 (54.13%) |
| **Trauma Exposure Factors** |  |
| Home Zones as on October 7 |  |
| Red | 48 (7.62%) |
| Orange | 181 (28.73%) |
| Yellow | 153 (24.29%) |
| Green | 248 (39.36%) |
| Know civilians murdered | 228 (36.19%) |
| Know soldiers murdered | 257 (40.79%) |
| Know persons taken hostage | 92 (14.60%) |
| Watch censored videos | 361 (57.30%) |
| Watch uncensored videos | 200 (31.75%) |
| **Obstetrics Factors** |  |
| Pregnant | 272 (43.17%) |
| Postpartum |  |
| Gave birth before October 7 | 135 (21.43%) |
| Gave birth after October 7 | 221 (35.08%) |

**Note.** Trauma exposure factors relate to the October 7 attacks that took place in Israel during the Israel-Hamas war. Home Zone refers to geographic zones as defined by the Israeli Defence Forces (IDF) Home Front Command to indicate the available time to reach shelter during missile attacks: Red ( ≤ 15 sec), Orange (30 sec), Yellow (45 sec), and Green (60-90 sec). History of mental illness refers to any mental illness diagnosed before October 7. Watched censored and
uncensored videos in the first two weeks; Values pertain to high frequency of exposure (score 5 or above on a 1–10 scale, 1 = not at all; 10 = many hours per day). N=630.

### 2.2 Measures

**Sociodemographic characteristics** (i.e., maternal age, education, marital status, religiosity, capturing degree of religiosity (i.e., Not at all to very religious), prior mental health illness, and obstetric information (i.e., obstetric status defined as (1) pregnant or postpartum at the time of assessment and (2) giving birth before or after October 7th) were collected using single-item measures.

Exposure to the October 7 attacks and war-related events concerned three forms of exposure. **Geographic exposure**, referring to trauma exposure defined by physical location and proximity to threat, was determined based on participants’ place of residence on October 7th and classified according to Israeli Defense Forces (IDF) Home Front Command alert zones, reflecting proximity to rocket and missile threats (Red = most proximate; Orange/Yellow = intermediate; Green = least proximate). **Interpersonal exposure** (indirect exposure), i.e., knowing someone affected, was assessed for personally knowing civilians or soldiers who were murdered or taken hostage during the attacks. **Media exposure**, i.e., exposure through news or social media, assessed engagement with October 7 attacks-related media, including censored and uncensored content, as well as watching videos circulated on digital platforms. Participants indicated the degree to which they engaged in each media form (censored, uncensored, watched, read) in the first 2 weeks following the attacks on a 10-point Likert scale (1 = not at all; 10 = many hours per day).

**Posttraumatic stress symptoms** were assessed using the PTSD Checklist for DSM-5 (PCL-5) (Weathers et al., 2018), a widely validated self-report measure of PTSD symptom severity. The PCL-5 consists of 20 items corresponding to DSM-5 PTSD symptoms experienced during the past month, each rated on a scale from 0 (“Not at all”) to 4 (“Extremely”), with higher scores indicating greater symptom severity. The PCL-5 is recommended by the U.S. Department of Veterans Affairs for provisional PTSD assessment, and a cutoff score of 33 or higher was used to indicate probable PTSD (Blevins et al., 2015; Weathers et al., 2018), and used in peripartum samples (Dekel et al., 2019). Internal consistency in the present sample was high (Cronbach’s α = 0.92).

**Resilience** was assessed using the Brief Resilience Scale (Smith et al., 2008) (BRS), a 6-item self-report measure designed to evaluate an individual’s perceived ability to recover from stress. Items are rated on a 5-point Likert scale from 1 (“strongly disagree”) to 5 (“strongly agree”), with higher scores reflecting greater resilience, conceptualized as the capacity to “bounce back” following stressful experiences. The BRS has demonstrated good reliability and construct validity across diverse populations (Smith et al., 2008; Windle et al., 2011) (α = 0.88).

**Social support was assessed** using the Multidimensional Scale of Perceived Social Support (MSPSS) (Zimet et al., 1988), a widely used 12-item self-report instrument measuring individuals’ perceptions of available social support. Items are rated on a 5-point Likert scale ranging from 1 (“very strongly disagree”) to 5 (“very strongly agree”), with higher scores indicating stronger perceived social support. The MSPSS has demonstrated excellent psychometric properties, including strong reliability and validity across diverse populations(Bedaso et al., 2021) (α = 0.93).

### 2.3 Data Preparation and Statistical Analyses

Data were checked for completeness prior to analysis. Participants with missing primary outcome data (i.e., PCL-5 scores) or invalid survey responses were excluded. Missing data were identified as missing at random and were handled (total missing is 1.16%) using multiple imputation with the *mice* package (Buuren and Groothuis-Oudshoorn, 2011) in R (version 4.3.2) employing predictive mean matching (pmm) to preserve observed distributions. Five imputed datasets (m = 5) were generated and pooled for inferential tests. The imputation model included demographic variables (i.e., age, marital status, education, prior mental-health illness, residential zone) and BRS and MSPSS items.

To compare rates of PTSD symptom cluster endorsement for the four DSM-5 clusters-intrusion, avoidance, negative alterations in cognition and mood, and alterations in arousal and reactivity-Cochran’s Q test was used with the DescTools package (Signorell A., 2023) for related proportions, followed by pairwise McNemar’s tests with Bonferroni correction for multiple comparisons. The figure was generated using the ggplot2 package in R (Wickham H., 2016).

To examine the contribution of media exposure to PTSD symptom severity, accounting for demographic background, psychosocial resources, and other forms of exposure, hierarchical multiple regression analysis was performed. Predictors were entered in four conceptual blocks: (1) demographics (i.e., age, marital status, education, prior mental health illness); (2) psychosocial variables (perceived resilience, perceived social support, religiosity); (3) geographical (residential zone) and interpersonal indirect exposure (acquaintance with victims/hostages); and (4) media exposure factors, entered last to assess their incremental contribution. Standardized beta values were generated using the *lm*.*beta* package(Behrendt S., 2023). Incremental R^2^ changes were examined to assess the additional variance explained at each step. Model assumptions (normality, multicollinearity, and outliers) were verified.

## 3. Results

Nearly 1 out of 4 participants (24.1%; n = 152) who were not directly exposed to the October 7 attacks met the clinical cutoff for probable PTSD regarding the attacks (PCL-5 total score >=33) around four months later. Rates of indirect exposure, knowing individuals murdered or taken hostage, were high ((Table 1).

### Prevalence of PTSD symptoms

Figure 1 presents the frequency of PTSD symptoms by DSM-5 symptoms cluster endorsement. Prevalence of PTSD symptom clusters differed significantly, Q(3) = 56.52, p < 0.001. Intrusive (62.7%) and alterations in arousal and reactivity (61.6%) symptoms were endorsed more frequently than avoidance symptoms (49.2%) and negative alterations in cognition and mood (52.4%) (all differences, p < 0.001). In contrast, no differences were observed between intrusive and alterations in arousal and reactivity symptoms or between avoidance and negative alterations in cognition and mood (both p = 1.00). Overall, intrusive and alterations in arousal and reactivity symptoms were the most frequently endorsed PTSD symptom clusters.

**Figure 1.**
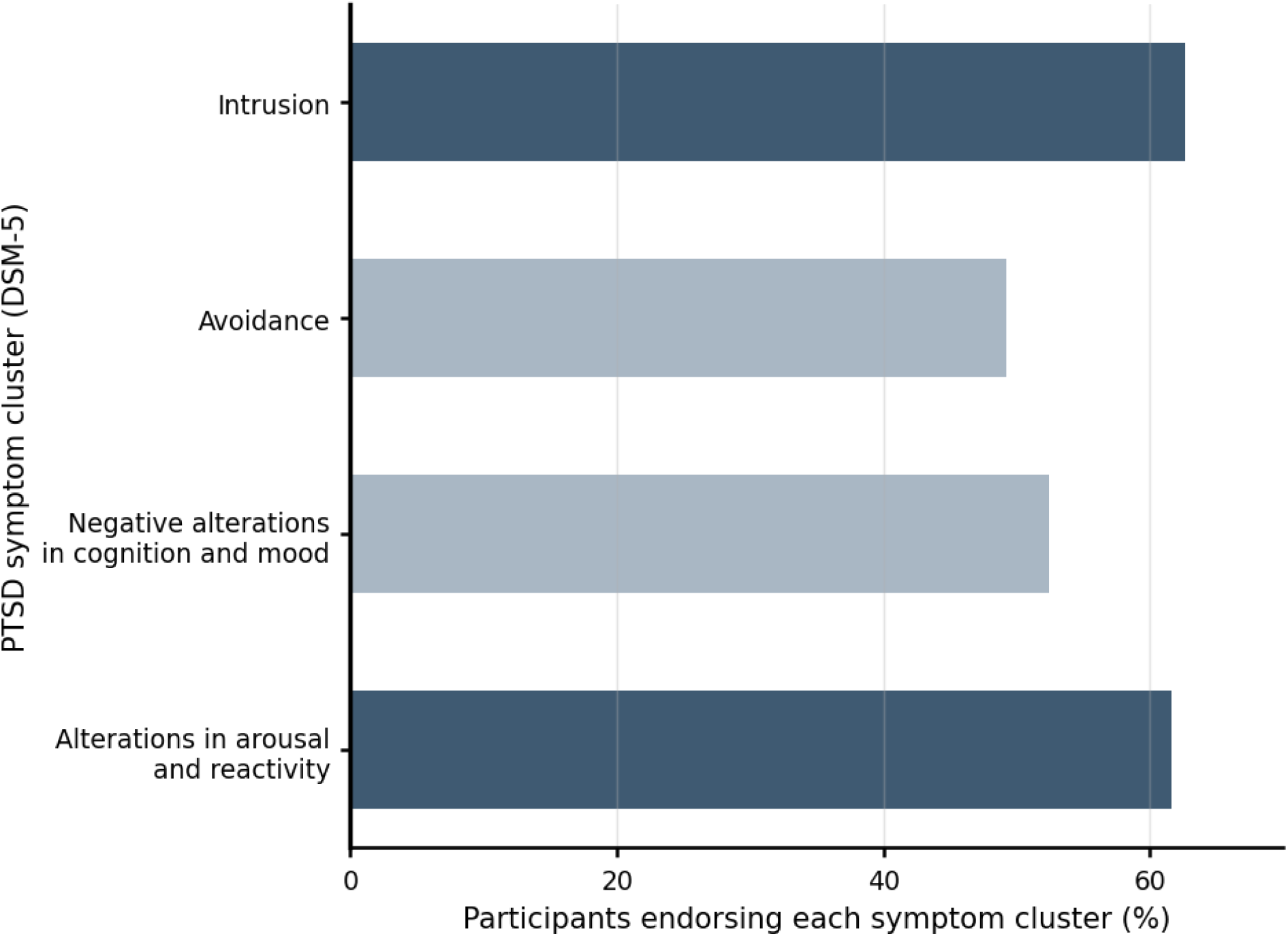
**Proportion of Participants meeting DSM-5** criteria for PTSD symptom cluster endorsement, assessed using the PCL-5. Intrusion was defined as endorsement of ≥1 item, whereas Hyperarousal, Negative Alterations in Cognition and Mood, and Avoidance required endorsement of ≥2 items. Items were considered endorsed if rated ≥2.

### Media exposure and PTSD symptoms

Table 2 presents hierarchical regression analysis for the prediction of PTSD. As can be seen, PTSD symptom severity was associated with prior mental health illness, psychosocial resources, direct and indirect exposure, and media exposure. Among the background factors, history of mental health illness (β = 0.105, p = 0.009) predicted higher PTSD symptom severity. Next, psychosocial resources collectively explained an additional 10.0% of the variance in symptoms (ΔR^2^ = 0.100). Less perceived resilience (β = -0.248, p < 0.001) and social support (β = -0.131, p < 0.001), and less religiosity (β = -0.130, p < 0.001) were each significantly associated with higher PTSD symptoms severity, Trauma exposure (i.e., geographic and interpersonal) variables accounting for an additional 2.3% of the variance (ΔR^2^ = 0.023) with indirect exposure pertaining to knowing civilians who were murdered or knowing individuals who were taken hostage each associated with higher symptom severity, Lastly, media exposure collective add a contribution of 2.8% of the variance (ΔR^2^ = 0.028). The degree of exposure to uncensored video content related to October 7 was significantly associated with higher PTSD severity (β = 0.153, p = 0.002), after accounting for demographics, psychosocial resources, and other forms of exposure (Table 2).

**Table 2.**
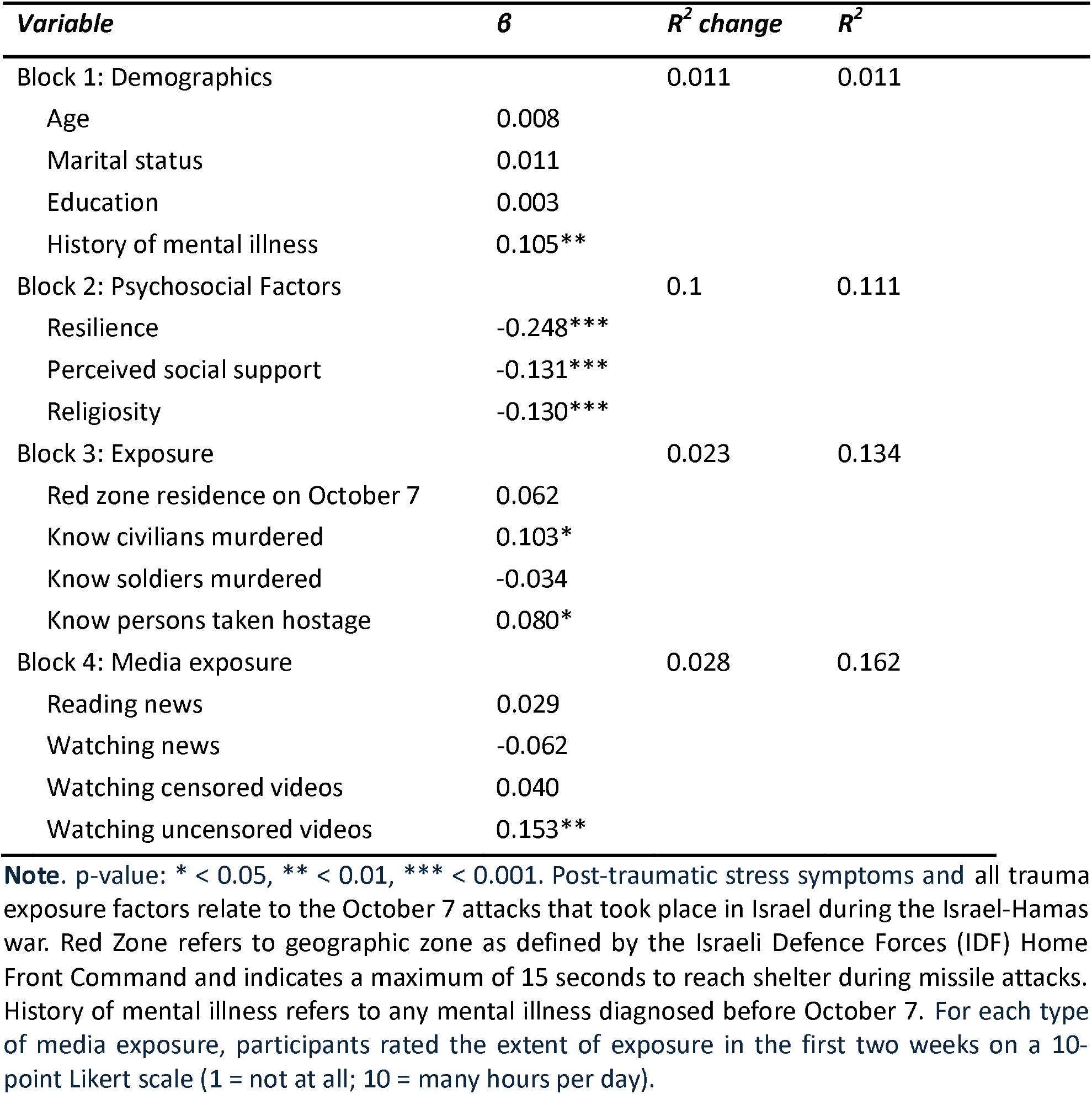
Hierarchical Regression Analysis of Demographic, Psychosocial, Exposure, and Media-Related Predictors of Post-Traumatic Stress Symptoms.

## 4. Discussion

Digital media have reshaped how individuals encounter traumatic events, extending their psychological impact well beyond direct exposure, yet the emotional consequences of this shift remain poorly understood. In this study, exposure to uncensored traumatic video content in the wake of a large-scale collective trauma was associated with greater PTSD symptom severity among non-directly exposed peripartum women, even after accounting for established risk factors, including sociodemographic, psychosocial resources, and other forms of trauma exposure. The findings suggest that real-time, graphic content may contribute to trauma-related symptomatology, beyond pathways traditionally described as indirect or secondary exposure (American Psychiatric Association, 2013).

Prior work on exposure to collective trauma via media and its effects on emotional coping has largely focused on traditional broadcast news, which is typically delayed, edited, and embedded within journalistic framing (Ahern et al., 2002; Garfin et al., 2020; Holman et al., 2014). The present findings extend this literature by providing new insight into the role of real-time, uncensored traumatic digital content in post-trauma adaptation. By distinguishing between exposure to uncensored digital material, censored video content, and traditional news media, our results indicate that the intensity and immediacy of uncensored video exposure may be particularly relevant for trauma-related symptom severity. Importantly, these associations remained after accounting for geographic proximity to threat, interpersonal exposure, sociodemographic factors, psychosocial resources, and baseline mental health, suggesting that the observed effects are not merely a reflection of greater objective exposure or pre-existing psychological distress. This pattern points to features intrinsic to contemporary digital environments, such as sensory vividness and temporal proximity (Holman et al., 2020), as potentially important mechanisms through which media exposure contributes to traumatic stress symptoms beyond overall media consumption volume.

Neurobiological and cognitive models of trauma offer a framework for understanding the heightened salience of uncensored video traumatic content. Experimental trauma-film paradigms indicate that viewing highly aversive visual material engages neural systems involved in threat processing and sensory memory encoding while limiting contextual and regulatory processing (Bourne et al., 2013; Clark et al., 2016). Unlike traditional media, uncensored digital videos often combine vivid visual imagery with distressing auditory cues-such as screams, explosions, or pleas for help-creating a multisensory experience that can strongly engage perceptual and emotional systems simultaneously. Even brief exposure to such material has been shown to alter memory-related neural connectivity, and neural responses to realistic aversive images predict subsequent PTSD symptom severity (Gvozdanovic et al., 2021; Portugal et al., 2023). These findings align with the revised dual representation theory of PTSD, which emphasizes the dominance of sensory-bound representations when contextual integration is constrained (Brewin, 2014; Brewin et al., 2010). Uncensored traumatic content, characterized by visual vividness, emotional intensity, and limited contextual framing, may therefore represent a particularly potent form of trauma exposure, even in the absence of direct physical exposure.

Consistent with the diathesis–stress model (McKeever and Huff, 2003), the findings indicate that trauma-related outcomes reflect the interplay between exposure severity and pre-existing individual vulnerability (Dekel et al., 2017; Grekin and O’Hara, 2014; Harville et al., 2010). A history of mental health illness and less perceived personal resilience and social support were associated with PTSD. As trait resilience and perceived social support may facilitate coping following trauma, it underscores the importance of securing support systems and enhancing resilience resources to buffer long-lasting psychological distress (Allouche-Kam et al., 2025; Bedaso et al., 2021; Smith et al., 2008; Zimet et al., 1988).

Clinically, the prominence of alterations in arousal and reactivity and intrusive symptoms observed in this study has important implications for understanding early posttraumatic responses in the context of digitally mediated collective trauma. Prior research indicates that these symptom domains commonly predominate in the acute aftermath of trauma and reflect heightened threat monitoring and involuntary re-experiencing processes (Bryant, 2011; Galatzer-Levy et al., 2018; Whitman et al., 2013). In this context, repeated exposure to uncensored, real-time, and visually vivid traumatic content may be particularly salient. Such exposure may sustain physiological arousal and reinforce threat-related attentional processes, while graphic visual material may increase the likelihood of intrusive recollections(Holman et al., 2014; Lamba et al., 2023; Silver et al., 2013). Consistent with this framework, exposure to uncensored traumatic content may preferentially amplify early-stage trauma responses, rather than later-emerging avoidance or cognitive symptoms. Together, these patterns underscore the potential value of early, trauma-informed screening and monitoring during periods of widespread exposure to uncensored graphic media, particularly among populations experiencing elevated baseline stress or vulnerability.

At a broader level, these findings foreground ethical and public-health considerations regarding population exposure to traumatic content during large-scale crises. The widespread availability of uncensored graphic material on digital platforms raises important questions about how to balance documentation and public awareness with psychological protection. Although the observed associations were modest in magnitude, their significance in the context of widespread and repeated exposure suggests potential relevance at the population level(Lamba et al., 2023; Yamin et al., 2025). From a public-health perspective, incorporating trauma-sensitive media practices and guidance around media exposure during crises may represent an important, and currently underdeveloped, avenue for mitigating exposure-related psychological burden.

This study has several strengths. We differentiated exposure to uncensored traumatic content from other forms of media exposure and assessed media exposure in a naturalistic, real-world context, reflecting how traumatic content is encountered during an unfolding collective trauma. Regression models simultaneously accounted for interpersonal, geographic, and media-based exposure, as well as established psychosocial risk factors, strengthening confidence that the observed associations were not attributable to broader exposure or pre-existing vulnerability alone.

Several limitations warrant consideration. Data were collected via an online self-report survey, a pragmatic approach during armed conflict that may be subject to reporting bias; however, self-reported exposure to traumatic events and media use is standard in large-scale studies of collective trauma, including those conducted following the September 11 attacks and the Boston Marathon bombings (Ahern et al., 2002; Holman et al., 2014). Retrospective reporting may introduce bias. Convenience sampling may limit generalizability, and the sample primarily reflects the Jewish Israeli population, raising the possibility of contextual specificity. In addition, the absence of diagnostic interviews limits conclusions regarding PTSD diagnoses, and the cross-sectional design precludes causal inference.

In summary, this study situates exposure to uncensored digital media within a broader framework of trauma-related risk, highlighting how features of contemporary media environments may shape early posttraumatic responses during collective crises. By integrating media exposure alongside interpersonal, geographic, and psychosocial factors, the findings suggest that digitally mediated exposure constitutes a distinct contextual dimension of trauma experience rather than a simple extension of traditional indirect exposure. This work underscores the need for future longitudinal research to clarify how repeated exposure to uncensored traumatic content interacts with individual vulnerability over time, and how such exposures may be addressed within trauma-informed public health and clinical response efforts.

## Data Availability

All data produced in the present study are available upon reasonable request to the authors

## Acknowledgements

The authors have no acknowledgements.

